# Urinary peptides predict future death

**DOI:** 10.1101/2023.04.28.23289257

**Authors:** Felix Keller, Joachim Beige, Justyna Siwy, Alexandre Mebazaa, Dewei An, Harald Mischak, Joost P. Schanstra, Marika Mokou, Paul Perco, Jan A. Staessen, Antonia Vlahou, Agnieszka Latosinska

**Author notes:** Corresponding author" Agnieszka Latosinska, PhD, Mosaiques-Diagnostics GmbH, Rotenburger Straße 20, 30659 Hannover, Germany.

## Abstract

**Background:** There is evidence of pre-established vulnerability in individuals that increases the risk of their progression to severe disease or death, though the mechanisms that cause this are still not fully understood. Previous research has demonstrated that a urinary peptide classifier (COV50) predicts disease progression and death from SARS-CoV-2 at an early stage, indicating that the outcome prediction may be partly due to already present vulnerabilities. The aim of this study is to examine the ability of COV50 to predict future non-COVID-19-related mortality, and evaluate whether the pre-established vulnerability can be generic and explained on a molecular level by urinary peptides.

**Methods:** Urinary proteomic data from 9193 patients (1719 patients sampled at intensive care unit (ICU) admission and 7474 patients with other diseases (non-ICU)) were extracted from the Human Urinary Proteome Database. The previously developed COV50 classifier, a urinary proteomics biomarker panel consisting of 50 peptides, was applied to all datasets. The association of COV50 scoring with mortality was evaluated.

**Results:** In the ICU group, an increase in the COV50 score of one unit resulted in a 20% higher relative risk of death (adj. HR 1·2 [95% CI 1·17-1·24]). The same increase in COV50 in non-ICU patients resulted in a higher relative risk of 61% (adj. HR 1·61 [95% CI 1·47-1·76]), in line with adjusted meta-analytic HR estimate of 1·55. The most notable and significant changes associated with future fatal events were reductions of specific collagen fragments, most of collagen alpha I(I).

**Conclusion:** The COV50 classifier is predictive of death in the absence of SARS-CoV-2 infection, suggesting that it detects pre-existing vulnerability. Prediction is based mainly on collagen fragments, possibly reflecting disturbances in the integrity of the extracellular matrix. These data may serve as basis for proteomics guided intervention aiming towards manipulating/improving collagen turnover, thereby reducing the risk of death.

## BACKGROUND

Pre-existing vulnerabilities have a key role in determining an individual’s risk for disease progression or death [1], highlighting the importance of considering these factors when managing diseases. Given the complexity of the disease-associated molecular mechanisms [2–5] and factors impacting the outcome, recognizing and understanding the pre-established vulnerabilities may help identifying high-risk individuals, tailoring treatment strategies, ultimately improving outcome.

Among theories attempting to explaining this phenomenon (e.g., in the context of trauma) [6], the “two-hit” model has evolved. Under this scenario, the stress response encompasses the physiological reaction to the initial injury (referred to as the “first hit”), followed by reaction to the secondary insult/ intervention (known as the “second hit”) [6, 7]. The model is rooted in the fundamental idea that consecutive insults, which may not have significant effects individually, can result in a profound physiological response. This reaction can manifest in various biological systems and can be evaluated by measuring multiple parameters [8]. However, the molecular mechanisms responsible for the “two-hit” model are complex and not fully understood [6, 8]. In general, the “first hit” acts as a priming event that predisposes the patient to develop systemic inflammatory syndrome, with a key feature being a leak of the endothelium, initially manifesting in a specific body region, but eventually affecting multiple organs. Subsequently, a second insult can trigger an exaggerated inflammatory response, responsible for potentially life-threatening conditions such as multiple organ failure and multiple organ dysfunction syndrome [6, 8]. Knowing the vulnerability to the “second hit” can support minimising the impact of complications, potentially leading to a better outcome.

Recently, it has been suggested that SARS-CoV-2 infection could act as a “second hit”. SARS-CoV-2 is among the main conditions associated with collapsing glomerulopathy, acting as a “second hit” in susceptible patients with APOL1 risk alleles, similarly to human immunodeficiency virus and other viruses [9]. Another example is complement-mediated disorder which seems to be a predominant form of thrombotic microangiopathy associated with COVID-19. Considering the development of thrombotic microangiopathy following SARS-CoV-2 infection, it was suggested that the virus acted as a “secondary trigger”, revealing an underlying complement defect [10].

Previous biomarker research demonstrated the capability of a urinary peptide-based classifier (COV50) to predict disease progression and death from SARS-CoV-2 at the earliest possible date, i.e., the first positive indication of a SARS-CoV-2 infection [11, 12]. The assessment was based on the measurement of 50 specific urinary peptides, with the most prominent changes being reduction of peptides derived from collagen alpha 1(I), polymeric immunoglobulin receptor and CD99 antigen, and an increase in peptides derived from alpha-1-antitrypsin [13]. The ability to predict outcome very soon after infection suggests that the prediction may not be solely based on molecular events associated with SARS-CoV-2 infection, but at least in part due to already present pre-established vulnerability to a “first hit”. This would indicate that prediction of severe disease course may be possible even before the infection. We hypothesised that this pre-established vulnerability can be generic (expanded to other indications) and COV50 could work act as a biomarker detecting vulnerable subjects, adversely affected by other clinical insults.

Hence, the present study aimed to examine the ability of COV50 to predict future non-COVID-19-related mortality in patients admitted to intensive care unit (ICU) or having other diseases (non-ICU). If the hypothesis is confirmed, a significantly higher number of this vulnerable population (defined by a COV50 score above the threshold) should experience death, compared to the population with a lower score.

## METHODS

### Patients

ICU: Patients from the medical, surgical, or mixed ICUs at 14 university hospitals from the FROG-ICU study were used [14]. Inclusion criteria were mechanical ventilation or administration of vasoactive agents for at least 24_h. The exclusion criteria were age under 18, severe head injury with a Glasgow Coma Scale below 8, brain death or persistent vegetative state, pregnancy or breastfeeding, transplantation in the past 12_months, moribund status, and lack of social security coverage. All capillary electrophoresis coupled to mass spectrometry (CE-MS) datasets where 1-year follow-up and information on relevant co-variables (age, body mass index (BMI), sex, blood pressure, estimated glomerular filtration rate (eGFR), presence of diabetes, kidney, cardiovascular disease, hypertension) was available were included in the present study without pre-selection.

Non-ICU: The assessment of COV50 in the non-ICU population was based on 7474 datasets from the Human Urinary Proteome Database [15, 16] with available information on age, sex, eGFR, blood pressure, BMI, presence of diabetes, kidney disease, cardiovascular disease, hypertension, and a follow-up data.

All datasets were from previously published studies and fully anonymized. Ethical review and approval were waived for this study by the ethics committee of the Hannover Medical School, Germany (no. 3116-2016), due to all data being fully anonymized. The number of subjects per study and patient characteristics are listed in **Table 1**.

**Table 1:** Descriptive statistics for the ICU and non-ICU samples analysed within this study.

|  | Level/Unit | ICU |  |  |  | Non-ICU |  |  |  |
| --- | --- | --- | --- | --- | --- | --- | --- | --- | --- |
|  |  | Overall | Death: no | Death: yes | p | Overall | Death: no | Death: yes | p |
| N |  | 1,719 (100%) | 1,139 (66,3%) | 580 (33,7%) |  | 7,474 (100%) | 6,849 (91,6%) | 625 (8,4%) |  |
| Study | FROG | 1,719 (100%) | 1,139 (100%) | 580 (100%) |  |  |  |  | <0,001 |
|  | CAD Predictions |  |  |  |  | 145 (1,9%) | 50 (0,7%) | 95 (15%) |  |
|  | CardioRen |  |  |  |  | 116 (1,6%) | 87 (1,3%) | 29 (4,6%) |  |
|  | DIRECT |  |  |  |  | 1,487 (20%) | 1,448 (21%) | 39 (6,2%) |  |
|  | EPOGH |  |  |  |  | 914 (12%) | 850 (12%) | 64 (10%) |  |
|  | EU Priority |  |  |  |  | 1,769 (24%) | 1,756 (26%) | 13 (2,1%) |  |
|  | GenScot |  |  |  |  | 473 (6,3%) | 417 (6,1%) | 56 (9,0%) |  |
|  | Heart Failure |  |  |  |  | 84 (1,1%) | 67 (1,0%) | 17 (2,7%) |  |
|  | Homage Fibrosis |  |  |  |  | 354 (4,7%) | 229 (3,3%) | 125 (20%) |  |
|  | PersTlgAN |  |  |  |  | 270 (3,6%) | 265 (3,9%) | 5 (0,8%) |  |
|  | Predictions |  |  |  |  | 91 (1,2%) | 85 (1,2%) | 6 (1,0%) |  |
|  | PROPHET |  |  |  |  | 462 (6,2%) | 444 (6,5%) | 18 (2,9%) |  |
|  | STOP IgAN |  |  |  |  | 109 (1,5%) | 107 (1,6%) | 2 (0,3%) |  |
|  | Sun Makro |  |  |  |  | 581 (7,8%) | 556 (8,1%) | 25 (4,0%) |  |
|  | TransBioBC |  |  |  |  | 131 (1,8%) | 117 (1,7%) | 14 (2,2%) |  |
|  | UZ Gent |  |  |  |  | 488 (6,5%) | 371 (5,4%) | 117 (19%) |  |
| Age | [yrs] | 62 (50, 73) | 58 (46, 69) | 70 (61, 78) | <0,001 | 60 (48, 68) | 59 (47, 66) | 73 (66, 79) | <0,001 |
| Female | yes | 602 (35%) | 414 (36%) | 188 (32%) | 0,11 | 2,857 (38%) | 2,657 (39%) | 200 (32%) | <0,001 |
| BMI | [kg/m<sup>2</sup>] | 26.2 (22.9, 30.0) | 26.2 (22.8, 29.9) | 26.4 (23.1, 30.1) | 0.6 | 27.5 (24.3, 31.2) | 27.6 (24.3, 31.4) | 26.9 (23.7, 30.1) | <0.001 |
| Systolic BP | [mmHg] | 123 (109, 140) | 124 (110, 140) | 120 (107, 139) | 0.01 | 132 (121, 145) | 132 (121, 144) | 138 (124, 153) | <0.001 |
| Diastolic BP | [mmHg] | 64 (55, 75) | 66 (56, 77) | 60 (52, 70) | <0.001 | 79 (72, 85) | 79 (73, 85) | 75 (67, 82) | <0.001 |
| Mean Arterial BP | [mmHg] | 84 (74, 95) | 86 (76, 96) | 80 (71, 92) | <0.001 | 97 (90, 104) | 97 (90, 104) | 97 (88, 105) | 0.3 |
| Hypertension | yes | 979 (57%) | 602 (53%) | 377 (65%) | <0.001 | 3,090 (41%) | 2,758 (40%) | 332 (53%) | <0.001 |
| eGFR | [ml/min/1.73 m<sup>2</sup>] | 87 (48, 127) | 97 (57, 132) | 67 (37, 107) | <0.001 | 82 (59, 99) | 84 (62, 100) | 61 (37, 80) | <0.001 |
| Kidney Disease | yes | 716 (42%) | 378 (33%) | 338 (58%) | <0.001 | 2,212 (30%) | 1,898 (28%) | 314 (50%) | <0.001 |
| Diabetes | yes | 280 (16%) | 160 (14%) | 120 (21%) | <0.001 | 4,101 (55%) | 3,938 (57%) | 163 (26%) | <0.001 |
| Cardiovascular Disease | yes | 98 (5.7%) | 49 (4.3%) | 49 (8.4%) | <0.001 | 1,357 (18%) | 983 (14%) | 374 (60%) | <0.001 |
| COV50 |  | 1.17 (0.34, 1.83) | 1.01 (0.11, 1.74) | 1.45 (0.72, 1.97) | <0.001 | -1.88 (-2.33, -1.27) | -1.89 (-2.34, -1.30) | -1.69 (-2.26, -0.94) | <0.001 |
| FU Duration | [month] | 12.0 (2.0, 12.1) | 12.0 (12.0, 12.5) | 0.7 (0.3, 2.1) | <0.001 | 47 (29, 67) | 48 (29, 67) | 38 (19, 62) | <0.001 |

Categorical variables are described with absolute (N) and group-wise relative frequencies (%), continuous variables with median (IQR). P-values for group differences result from chi-squared homogeneity tests for categorical and for Wilcoxon rank sum test for continuous variables. **Abbreviations:** BMI= body mass index, BP= blood pressure, eGFR= estimated glomerular filtration rate, FU= follow-up, ICU= intensive care unit, yrs= years

### Urinary proteome/ peptidome data

The urinary proteome is well characterized and reference standards are available [17]. Urinary proteome analysis was performed on urine samples collected at study inclusion which were bio-banked until assayed. Detailed information on urine sample preparation, proteome analysis by CE-MS, data processing, and sequencing of the urinary peptides allowing identification of parental proteins is available in previous publications [18–20].

### Outcome

In the FROG-ICU study, information on vital status was collected 3, 6, and 12_months after ICU discharge, as previously described [21]. For the non-ICU patients, vital status and outcome was assessed as described in the specific original studies [18, 22–37].

### Statistics

As descriptive statistics for the ICU and non-ICU samples, shown in **Table 1**, median and 1^st^ and 3^rd^ quartile (IQR) were used for continuous variables and absolute (N) and relative frequencies (%) for categorical variables. Hypotheses of no differences in scale or distribution of patient characteristics between the death and non-death groups were tested with Wilcoxon–Mann–Whitney tests for continuous and with χ^2^-homogeneity tests for categorical variables.

Kernel density estimates of the distribution of COV50 scores split by ICU and mortality groups are displayed in **Figure 1 A**. Mortality per person-time stratified by age and COV50 groups, seen in **Figure 1 B**, is estimated as the ratio of the number of the deceased to the sum of all patients’ observation times within each group scaled to 100 person-years. The corresponding mortality probabilities and their 95% confidence intervals (CI) for each group, as seen in **Figure 1 C**, represent estimates from a logistic regression including all 9193 patients.

**Figure 1:**
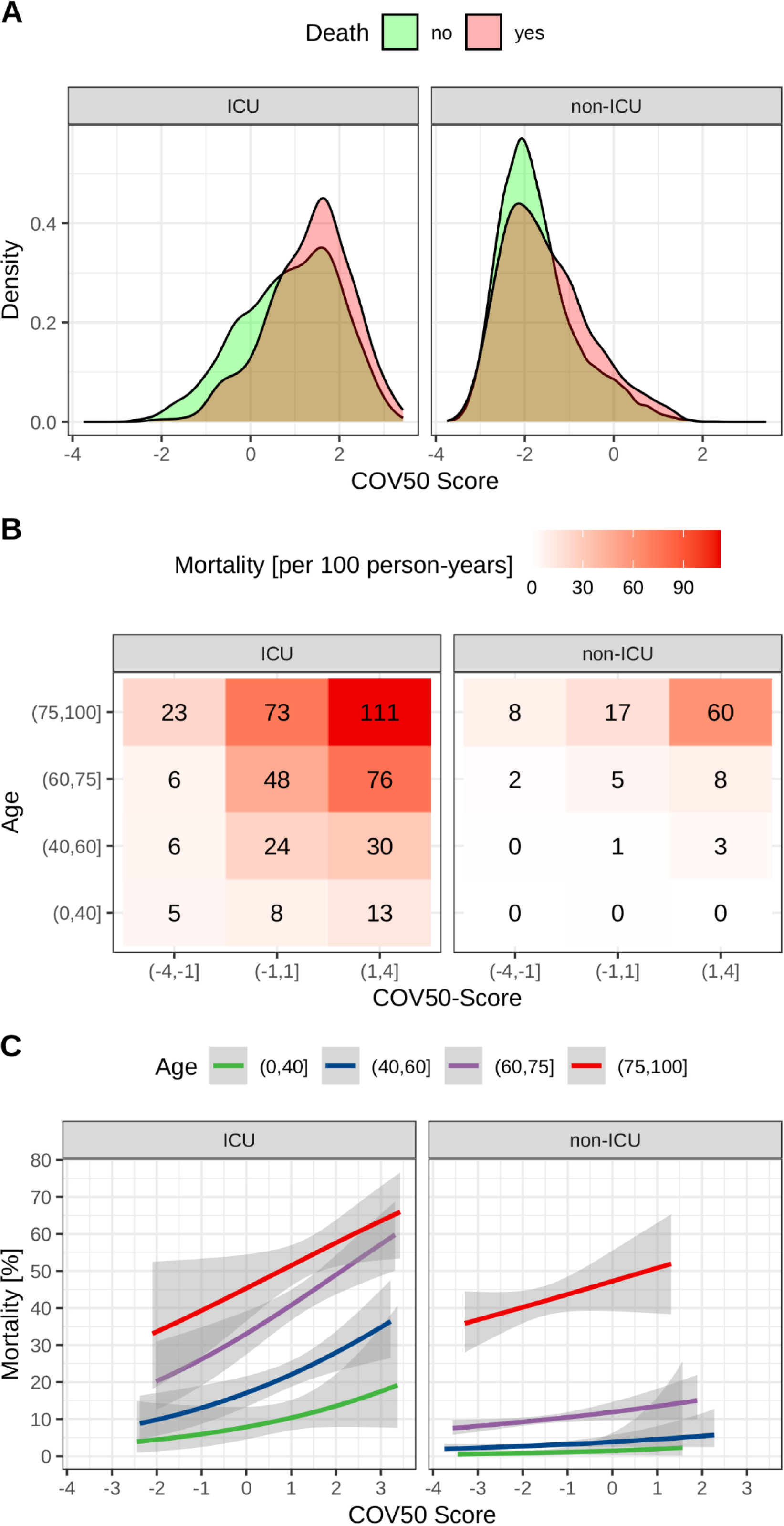
A) Density of the COV50 distribution in ICU and non-ICU subjects. B) Mortality per person-years for FROG and non-ICU cohorts given age and COV50. C) Mortality as share [0–1] from a logistic regression for FROG and non-ICU cohorts given age and COV50.

For each study, separate unadjusted Cox regressions for the effect of the COV50 score on experiencing death were performed, as listed in **Figure 2 A**. In **Figure 2 B** these models were additionally adjusted for age, female, log(BMI), mean arterial pressure (MAP) and log(eGFR). All regressors besides female and COV50 were normalized (mean 0, sd 1). The natural logarithms of the estimated hazard ratios (logHR) and their standard errors were combined in meta-analyses to determine the effect of the COV50 score on mortality. A random effects model was estimated after the assumption that all included studies are heterogeneous, i.e., coming from different populations. Study weights are based on the logHR estimates’ uncertainty, namely their standard errors. Studies were categorized into more homogenous subgroups, and estimates for each subgroup are displayed in **Figure 2**. Overall and group-wise between-study heterogeneity is presented with τ^2^ and assessed by Higgins & Thompson’s I^2^ statistic. χ^2^-Tests for heterogeneity and subgroup differences are based on Cochran’s Q. Random effects meta-analysis estimates are presented with 95% CIs and a 95% prediction interval for the overall effect is given. One Cox regression stratified by study pooling all 9193 patients was used as a benchmark to the meta-analytic approach. As displayed in **Table 2**, the model’s adjustment specification is identical to the adjusted separate study regressions (**Figure 2 B**). To be comparable to the adjusted meta-analytic estimate, HRs for COV50 interacted with ICU and non-ICU, as well as for the above-mentioned non-ICU subgroups, were estimated. Standard errors were clustered on the study level for more robust inference and due to unobserved heterogeneity between studies. The models log-likelihood, it’s Wald test and concordance are reported in **Table 2**. We allow for a type 1 error of 5%, all hypotheses are two-sided. All analyses were carried out using R 4.2.2.

**Figure 2:**
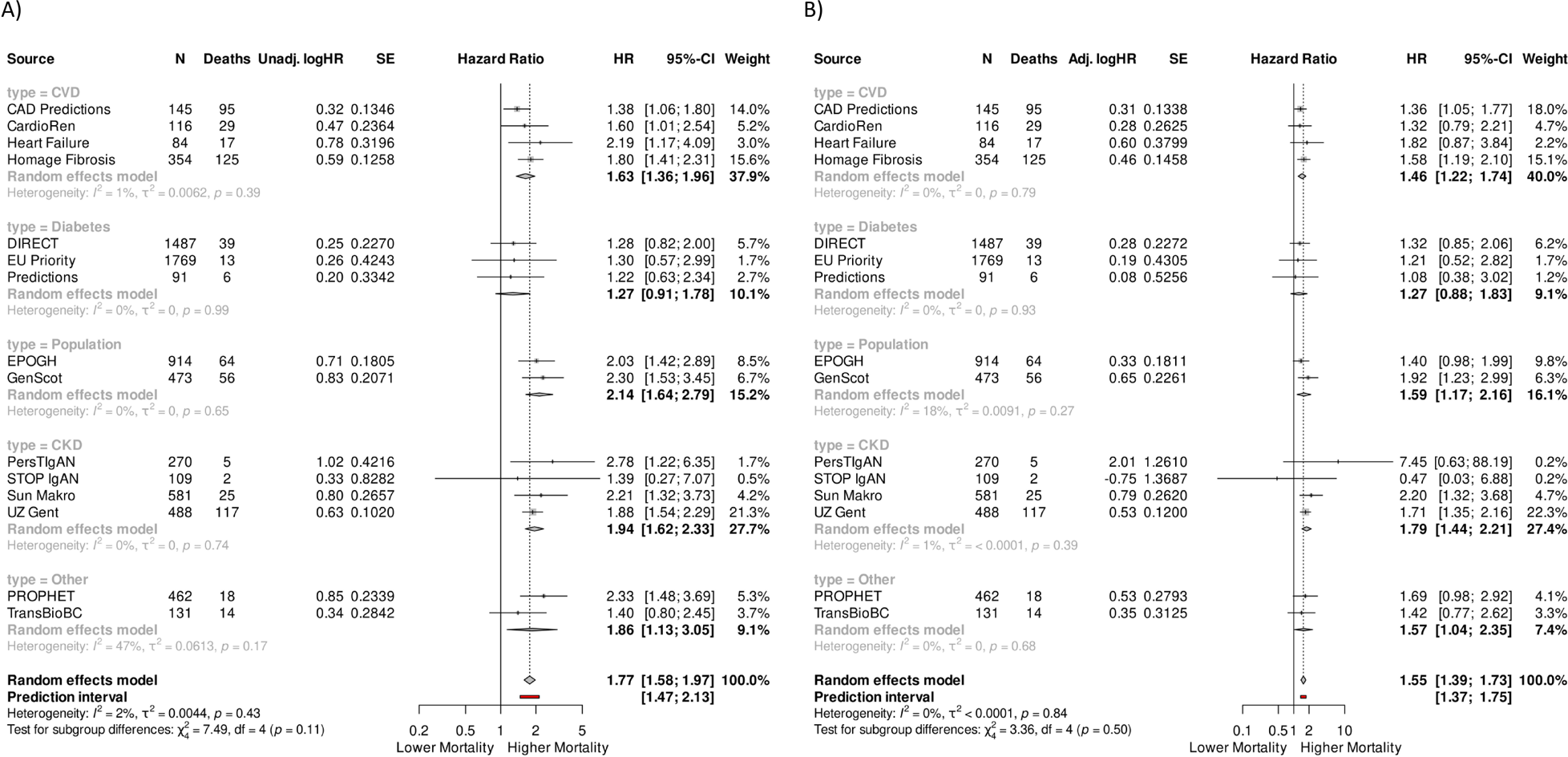
Random effects meta analyes based on the log-HR and the standard errors from the separate cox regressions. A) Unadjusted, B), adjusted for sex, age, kidney function and BMI. The size of dot symbols is proportional to weight and weight is inverse proportional to HR standard error.

**Table 2:** Estimates from the pooled adjusted Cox regression.

| Effect | Group | (Non-ICU Subgroup) | HR | 95% CI | p |
| --- | --- | --- | --- | --- | --- |
| Age |  |  | 1.83 | 1.43, 2.35 | <0.001 |
| Female |  |  | 0.78 | 0.69, 0.87 | <0.001 |
| log(BMI) |  |  | 0.91 | 0.87, 0.94 | <0.001 |
| MAP |  |  | 0.88 | 0.84, 0.93 | <0.001 |
| log(eGFR) |  |  | 0.89 | 0.84, 0.95 | <0.001 |
| COV50 | ICU |  | 1.2 | 1.17, 1.24 | <0.001 |
|  | Non-ICU |  | 1.61 | 1.47, 1.76 | <0.001 |
|  |  | CVD | 1.51 | 1.36, 1.68 | <0.001 |
|  |  | Diabetes | 1.24 | 1.23, 1.25 | <0.001 |
|  |  | Population | 1.68 | 1.31, 2.15 | <0.001 |
|  |  | CKD | 1.82 | 1.70, 1.94 | <0.001 |
|  |  | Other | 1.75 | 1.45, 2.11 | <0.001 |

**Abbreviations:** BMI = body mass index, CI = Confidence Interval, CKD = chronic kidney disease, eGFR = estimated glomerular filtration rate, HR = Hazard Ratio, ICU = intensive care unit; MAP= mean arterial pressure.
All regressors besides Female and COV50 are were normalized to mean 0 and sd 1.
n = 9,193; N events = 1,205;
statistic.wald = 7,096; p = <0.001;
c-index = 0.618; c-index SE = 0.013;
Log-likelihood = -5,959;

## RESULTS

First, we evaluated the hypothesis that the COV50 classifier defines a vulnerable population at the molecular level, irrespective of SARS-CoV-2 infection. For that purpose, we investigated datasets from subjects from the FROG-ICU study [21], as this study was more comparable to the CRIT-COV study (patients in ICU) and had available a large number of endpoints. We identified 1719 datasets to be included in this study, for which follow-up and information on relevant co-variables were available [38].

To further support our analysis, we also investigated if COV50 enables death prediction in subjects, not in ICU. Studies with more than 50 individuals and available follow-up information were selected from the Human Urinary Proteome Database [15, 16], applying the same criteria as above regarding the availability of co-variables.

The demographic information on the subjects included in the study split by ICU and death is given in **Table 1**. Among the risk factors for death we found significant differences at the aggregate level in both, the ICU and the non-ICU subgroups, as expected. The median COV50 score is significantly (p <0·001) higher in patients that were to experience death during the observation time, as also displayed in its distributions in Panel A of **Figure 1**.

As age is a crucial risk factor for death, we investigated the relationship between COV50 and mortality split by age groups. The results are shown in panels B and C of **Figure 1**. Panel B describes mortality in person-time in COV50 groups, whereas panel C relates mortality as a percentage with continuous COV50. In both subgroups, an increase in COV50 accompanies higher mortality, with the effect being more pronounced at higher age.

Crude HRs in **Figure 2 A** for all studies in general show an association of higher relative risk of death with increasing COV50 scores, with all but 5 studies showing a significantly elevated relative risk. For all subgroups in the meta-analysis, besides the diabetes-related studies, the combined estimate for the HR is significantly different from 1, as indicated by the 95% CIs. In general, adjustment for risk factors lowered the COV50 HR estimates, which aligns with expectations, as adjustment generally improves comparability by accounting for observed between study heterogeneity on the patient level. However, in studies (particularly PersTIgAN, STOPIgAN) with low numbers of events, variance increased drastically with the adjustment. The estimates from the meta-analysis resulted in an unadjusted HR of 1·77 [95% CI 1·58-1·97] and an adjusted HR of 1·55 [95% CI 1·39-1·73]. Though appearing on a trend level within and between the subgroups, neither heterogeneity nor subgroup differences were significant (**Figure 2**).

Subgroup HR estimates from the adjusted meta-analysis in **Figure 2 B** are robust, since they are close to corresponding estimates from the pooled adjusted Cox regression in **Table 2**. In the ICU group, an increase in the COV50 score of one unit results (on average, ceteris paribus) in a 20% higher relative risk of death (adj. HR 1·2 [95% CI 1·17-1·24]). As the absolute risk of death is considerably lower in non-ICU patients, the same increase in COV50 in non-ICU patients results in a higher relative risk of 61% (adj. HR 1·61 [95% CI 1·47-1·76]). This is in line with the adjusted HR estimate of 1·55 from the meta-analysis.

COV50 is a composite score based on 50 distinct urinary peptides. To examine which of these 50 peptides served as individual predictors of death in the cohorts investigated (ICU, non-ICU), we compared the distribution of the 50 peptides in the datasets from survivors with those from subjects that died. The outcome was assessed at 1 and 5 years for ICU and non-ICU cohorts, respectively. The results of this analysis are shown in **Table 3**. A high degree of concordance was found when comparing the peptides regulation trend in the context of COVID-19, death in or after ICU, or death without ICU stay. The most notable and significant changes were associated with future fatal events are the reductions of specific collagen fragments, most of collagen alpha I(I).

**Table 3:**
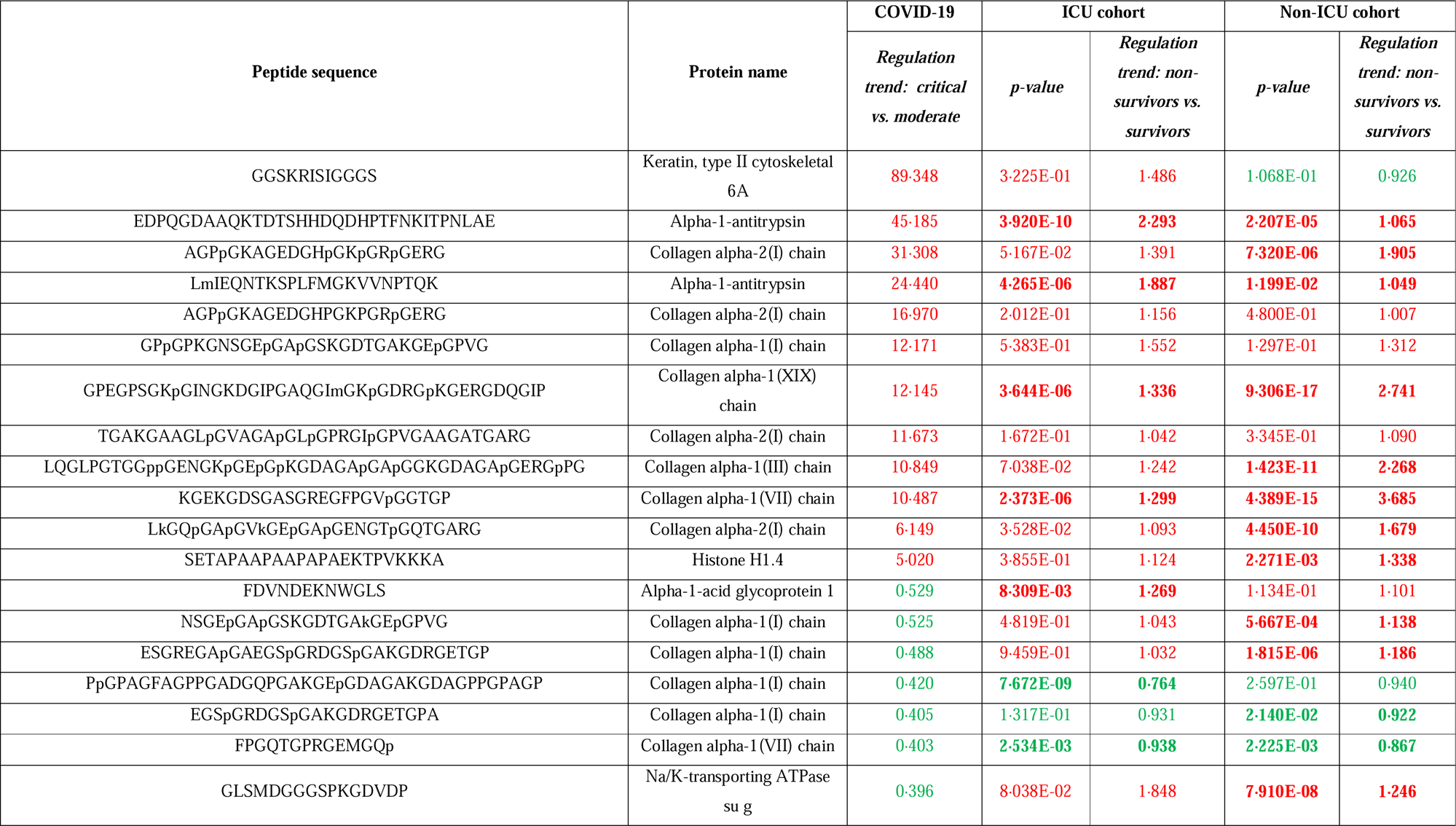
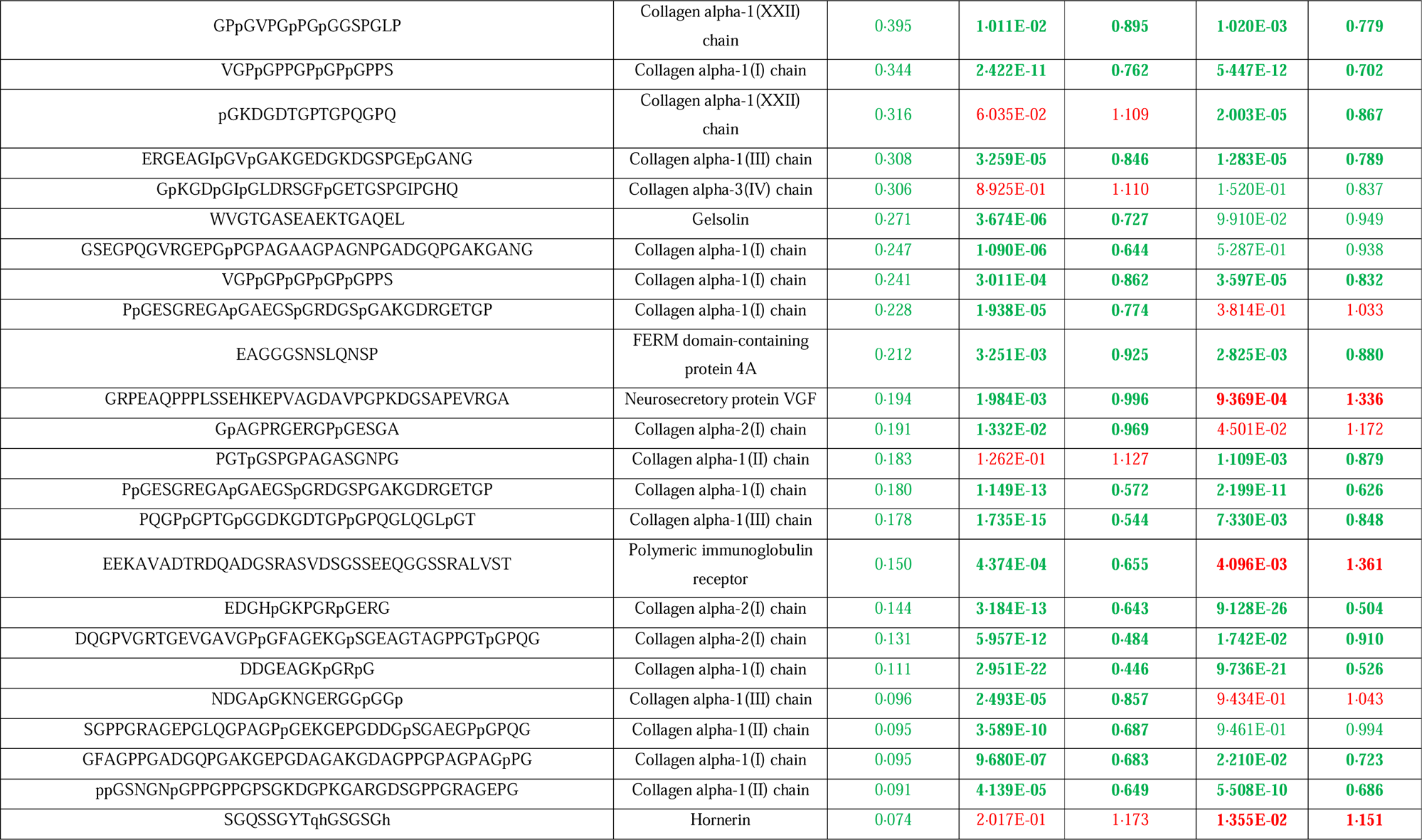
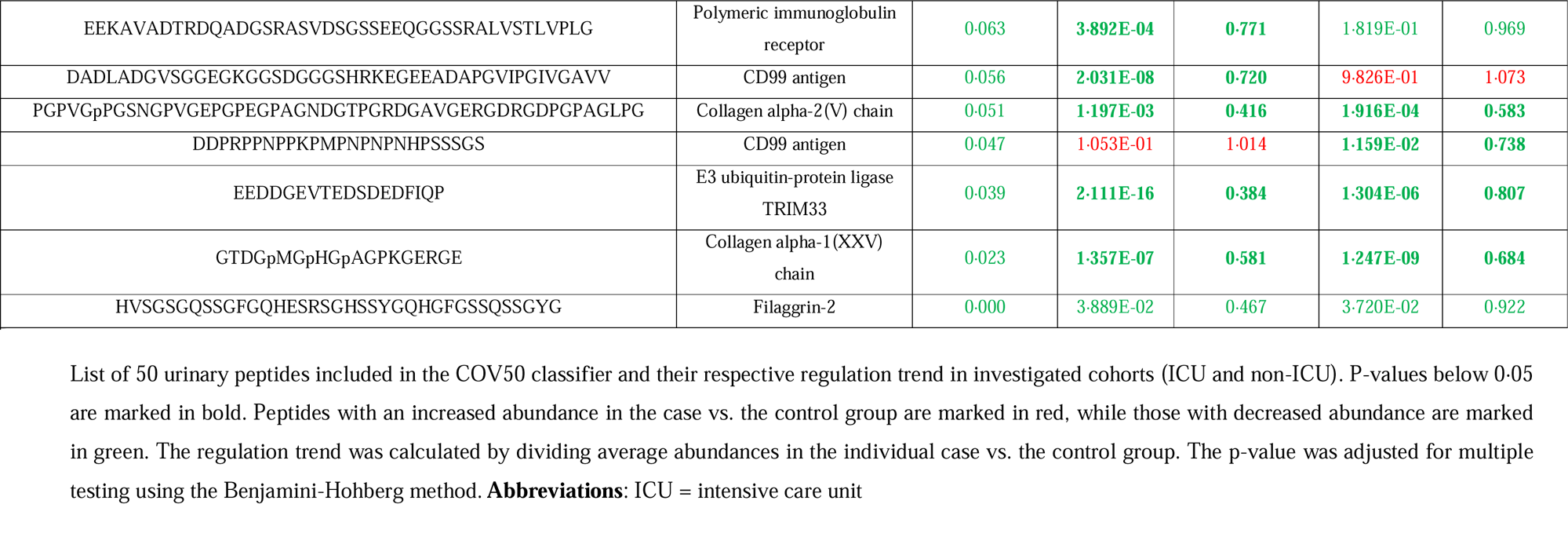
Urinary peptides included in the COV50 classifier.

In the ICU subjects, 28 of the 50 peptides were found to be significantly associated with future death. Of these, 26 showed a regulation trend in a similar direction as for critical/lethal COVID-19. At the same time, two peptides, one from alpha-1-acid glycoprotein 1 and one from sodium/potassium-transporting ATPase subunit gamma, had an opposing regulation. When investigating the most prominent peptides derived from collagen, all significant changes are concordant between death in COVID-19 or ICU. In the non-ICU subjects, 36 of the 50 peptides were significantly associated with future death. Of these, 27 showed a regulation concordant with the one in critical/lethal COVID-19, while 9 peptides changed in an opposing direction. The latter are the peptides derived from alpha-1-acid glycoprotein 1, polymeric immunoglobulin receptor, neurosecretory protein VGF, CD99 antigen, hornerin, collagen alpha-1(I), and collagen alpha-2(I).

The most prominent difference in comparison to the distribution in COVID-19 patients was observed for peptides derived from CD99 and polymeric immunoglobulin receptor. While in critical COVID-19 patients a consistent and significant reduction of multiple CD99 and polymeric immunoglobulin receptor peptides was associated with severe disease and mortality, this was not observed in ICU and non-ICU populations not infected with SARS-CoV2.

## DISCUSSION

This study is the first to investigate a peptide-based classifier, COV50, and specific urinary peptides in a large and diverse population of patients in and outside the ICU. The data demonstrate that COV50 not only predicts an unfavourable outcome of a COVID-19 episode but apparently identifies “vulnerable subjects”, who are likely at substantially higher risk of severe or lethal COVID-19. This vulnerability also appears relevant in other clinical situations (e.g., non-SARS-CoV-2 infections) including those leading to ICU admission, increasing the risk of death in most pathological conditions. This concept of the “second hit” phenomenon exaggerated on the grounds of a pre-established condition (“vulnerability”), and inappropriate innate immune response to injury and trauma has been related to perfusion changes of critical compartments and pathological sequelae [6].

The most prominent and consistent findings are the reduction of several specific urinary collagen fragments, most from collagen alpha-I(I). The reduction of these collagen fragments may indicate reduced collagen degradation in the extracellular matrix, which is expected to result in increased fibrosis. Fibrosis has been associated with various diseases affecting different organs, including the liver, kidney, lungs, and heart [39]. Previous studies revealed association of fibrosis with poor outcome in patients with different pathologies [40–42]. Fibrosis may constitute the “first hit” and induce vulnerability to “second hit” events either in e.g. infectious or general (cardiovascular) scenario. In this case, a pre-existing fibrotic condition may make an organ/ tissue more susceptible to further damage or insults from a second event or trigger. Fibrosis alters the normal structure and function of the affected tissue, compromising its ability to respond and recover from subsequent insults. Consequently, when a second hit, such as infection or inflammation occurs, it can lead to more severe complications and worsen the overall outcome.

The concordance of significant changes in individual peptides observed due to critical/lethal COVID-19 appears to be higher in the context of ICU than in non-ICU subjects. While an objective measure to assess significant differences does not seem to exist, a concordance (based on up- or down-regulation) of 93% (in the case of ICU) compared to 75% (in the case of non-ICU) is at least indicative.

As expected, similarities between changes in biomarkers in patients developing the critical condition, irrespective of the underlying pathology and disease aetiology, could be observed. At the same time it becomes evident that specific changes, a decrease of peptides from CD99 antigen and polymeric immunoglobulin receptor, are associated explicitly with critical COVID-19, and cannot be associated with all-cause death, neither in nor outside ICU. Thus, it is reasonable to assume that the “second hit” in the context of a SARS-CoV-2 infection is depicted via peptides deregulated in severe COVID-19 only, like CD99 antigen and polymeric immunoglobulin receptor.

This study’s findings agree with previous research that reported an association of urinary peptides (or classifiers based on these) with unfavourable outcome. A search in Pubmed for the keywords (urine OR urinary) AND (peptidom* OR proteom*) AND (death OR mortality) in the title or abstract resulted in 96 publications. Upon manual inspection by three authors, 11 manuscripts were found to be relevant, describing studies investigating the association of urinary peptides with mortality in humans, including the manuscripts describing the development of COV50 [11, 12]. Currie et al. described a significant value of CKD273, a classifier based on 273 urinary peptides, in predicting mortality in 155 microalbuminuric type 2 diabetic patients [43]. Similar results were also presented by Verbeke et al., where CKD273 was significantly associated with mortality in 451 chronic kidney disease patients [36]. Nkuipou-Kenfack et al. reported the association of urinary peptides with death, and the development of a classifier based on these to predict mortality after ICU stay in 1243 patients [38]. In 2021, Martens et al. described the association of multiple urinary peptides, many of these collagen fragments, with biological age, and mortality [18]. Batra et al. presented a proteomics-based mortality signature in COVID-19 and acute respiratory distress syndrome patients [44]. In the context of hepatocellular carcinoma, Bannaga et al. described several urine peptides being significantly associated with death [45]. Very recently, Wei et al. reported on the detection of urinary peptides related to pulse-wave velocity also being associated with mortality [46]. In a very well-powered study on 1170 patients that underwent cardiac surgery, Piedrafita and colleagues reported the identification of 204 urinary peptides associated with acute kidney injury [47]. A classifier based on these 204 peptides was validated in an independent cohort of 1569 ICU patients, demonstrating good performance and significant association with mortality. In almost all of these studies, collagen peptides were among the most prominent biomarkers, with reduced abundance being associated with increased risk of death, as also demonstrated recently by He and colleagues in the context of heart failure [25]. The data on quite large cohorts in ICU and subjects not in critical condition at the time of sampling indicate that urinary peptides and classifiers derived thereof hold significant predictive value for a patient-relevant endpoint: death. In complete agreement with previous studies, prediction of death appears to be based mainly on collagen fragments, which may indicate attenuation of collagen degradation, consequently progressing fibrotic processes. Evidently, the COV50 classifier was not developed to predict death in the general population. Also, based on the observation in this study that several peptides contained in this classifier show opposite regulation on predicting critical COVID-19 or death from any cause, it is to be expected that a classifier developed explicitly for prediction of death, based on only those peptides significantly associated with death, could be of substantial value in guiding death-preventing interventions. It is to be expected that such classifier would be based mainly on collagen fragments.

The study has limitations. It has been performed based on previously generated datasets. However, the large number of datasets, the high number of endpoints assessed, and the very high significance level of the findings strongly support that the results can be generalized. Along these lines, a strength of the study is the inclusion of datasets from multiple studies, indicating a robust basis for the assessment.

## CONCLUSIONS

Collectively, in this study we demonstrate that the urinary COV50 classifier is significantly associated with future death in ICU patients as well as in non-ICU patients, and enables the detection of “vulnerable” subjects, irrespectively of the underlying conditions. Further research is needed to assess if specific, personalized intervention guided by urinary collagen fragments can significantly improve outcomes, reducing future death.

## Data Availability

Anonymised data and code used in conducting the analyses will be made available upon request directed to the corresponding author. Proposals will be reviewed and approved by the authors with scientific merit and feasibility as the criteria. After approval of a proposal, data can be shared via a secure online platform after signing a data access and confidentiality agreement. Data will be made available for a maximum of 5 years after a data sharing agreement has been signed

## LIST OF ABBREVIATIONS

CE-MS: capillary electrophoresis coupled to mass spectrometry
ICU: intensive care unit
BMI: body mass index
eGFR: estimated glomerular filtration rate
HR: hazard ratios

## DECLARATIONS

### Ethical approval and consent to participate

All datasets were from previously published studies and fully anonymized. Ethical review and approval were waived for this study by the ethics committee of the Hannover Medical School, Germany (no. 3116-2016), due to all data being fully anonymized.

### Consent for publication

Not applicable

### Competing interests

HM is the cofounder and co-owner of Mosaiques Diagnostics (Hannover, Germany) and AL, MM, and JS are employees of Mosaiques Diagnostics. PP is also employed by Delta4 GmbH. AM reports grants or contracts from 4TEEN4, Abbott, Roche and Sphyngotec, and consulting fees from Roche, Adrenomed, Corteria, Fire1 and payment or honoraria from Merc and Novartis. All other authors declare no competing interests.

### Funding

This work was supported in part by funding through the European Union’s Horizon Europe Marie Skłodowska-Curie Actions Doctoral Networks - Industrial Doctorates Programme (HORIZON – MSCA – 2021 – DN-ID, “DisCo-I”, grant No 101072828) to AL, AV, JPS and HM, and in part by BMBF via the “Propersis” project, grant No 01DN21014 to HM. The funders had no role in the design of the study; collection, analyses, or interpretation of data; in the writing of the manuscript; or in the decision to publish the results.

### Authors’ contributions

AL, HM, AV, JS and FK conceptualised the study. JS, AL, MM, JPS and HM collected and curated the urine proteomic data. AL, FK, PP and HM performed the statistical analysis. JB, AM, JAS and DA provided access to the patient data. JB and JAS had access to and verified the data reported in this study. AL, HM and FK wrote the first draft of the manuscript. All authors interpreted the results, commented on and revised successive drafts of the manuscript, and approved the final version. All authors had full access to all the data in the study and had final responsibility for the decision to submit for publication.

## Acknowledgments

All authors are grateful to all patients who contributed samples. The STOP-IgAN trial was conducted by the RWTH Aachen University (T. Rauen, J. Floege) and funded by the German Federal Ministry of Education and Research (GFVT01044604).

## Notes

### Funding Statement

This work was supported in part by funding through the European Union Horizon Europe Marie Sklodowska-Curie Actions Doctoral Networks - Industrial Doctorates Programme (HORIZON-MSCA-2021-DN-ID, DisCo-I, grant No 101072828) to AL, AV, JPS and HM, and in part by BMBF via the Propersis project, grant No 01DN21014 to HM. The funders had no role in the design of the study; collection, analyses, or interpretation of data; in the writing of the manuscript; or in the decision to publish the results.

### Summary of Updates

The abstract, introduction, and discussion have been revised, aiming to increase the article's clarity.

## REFERENCES

1. Gallo Marin B, Aghagoli G, Lavine K, Yang L, Siff EJ, Chiang SS, et al. Predictors of COVID-19 severity: A literature review. Rev Med Virol. 2021;31(1):1–10.

2. Hartog N, Faber W, Frisch A, Bauss J, Bupp CP, Rajasekaran S, et al. SARS-CoV-2 infection: molecular mechanisms of severe outcomes to suggest therapeutics. Expert Rev Proteomics. 2021;18(2):105–18.

3. Hojyo S, Uchida M, Tanaka K, Hasebe R, Tanaka Y, Murakami M, et al. How COVID-19 induces cytokine storm with high mortality. Inflamm Regen. 2020;40:37.

4. Li CX, Gao J, Zhang Z, Chen L, Li X, Zhou M, et al. Multiomics integration-based molecular characterizations of COVID-19. Brief Bioinform. 2022;23(1).

5. Tay MZ, Poh CM, Renia L, MacAry PA, Ng LFP. The trinity of COVID-19: immunity, inflammation and intervention. Nat Rev Immunol. 2020;20(6):363–74.

6. Pape HC, Moore EE, McKinley T, Sauaia A. Pathophysiology in patients with polytrauma. Injury. 2022;53(7):2400–12.

7. Moore FA, Moore EE. Evolving concepts in the pathogenesis of postinjury multiple organ failure. Surg Clin North Am. 1995;75(2):257–77.

8. Morris CF, Tahir M, Arshid S, Castro MS, Fontes W. Reconciling the IPC and Two-Hit Models: Dissecting the Underlying Cellular and Molecular Mechanisms of Two Seemingly Opposing Frameworks. J Immunol Res. 2015;2015:697193.

9. Velez JCQ, Caza T, Larsen CP. COVAN is the new HIVAN: the re-emergence of collapsing glomerulopathy with COVID-19. Nat Rev Nephrol. 2020;16(10):565–7.

10. Tiwari NR, Phatak S, Sharma VR, Agarwal SK. COVID-19 and thrombotic microangiopathies. Thromb Res. 2021;202:191–8.

11. Staessen JA, Wendt R, Yu YL, Kalbitz S, Thijs L, Siwy J, et al. Predictive performance and clinical application of COV50, a urinary proteomic biomarker in early COVID-19 infection: a prospective multicentre cohort study. Lancet Digit Health. 2022;4(10):e727–e37.

12. Wendt R, Thijs L, Kalbitz S, Mischak H, Siwy J, Raad J, et al. A urinary peptidomic profile predicts outcome in SARS-CoV-2-infected patients. EClinicalMedicine. 2021;36:100883.

13. Siwy J, Wendt R, Albalat A, He T, Mischak H, Mullen W, et al. CD99 and polymeric immunoglobulin receptor peptides deregulation in critical COVID-19: A potential link to molecular pathophysiology? Proteomics. 2021;21(20):e2100133.

14. Gayat E, Cariou A, Deye N, Vieillard-Baron A, Jaber S, Damoisel C, et al. Determinants of long-term outcome in ICU survivors: results from the FROG-ICU study. Crit Care. 2018;22(1):8.

15. Latosinska A, Siwy J, Mischak H, Frantzi M. Peptidomics and proteomics based on CE-MS as a robust tool in clinical application: The past, the present, and the future. Electrophoresis. 2019;40(18-19):2294–308.

16. Rodriguez-Suarez E, Siwy J, Zurbig P, Mischak H. Urine as a source for clinical proteome analysis: from discovery to clinical application. Biochim Biophys Acta. 2014;1844(5):884–98.

17. Mischak H, Kolch W, Aivaliotis M, Bouyssie D, Court M, Dihazi H, et al. Comprehensive human urine standards for comparability and standardization in clinical proteome analysis. Proteomics Clin Appl. 2010;4(4):464–78.

18. Martens DS, Thijs L, Latosinska A, Trenson S, Siwy J, Zhang ZY, et al. Urinary peptidomic profiles to address age-related disabilities: a prospective population study. Lancet Healthy Longev. 2021;2(11):e690–e703.

19. Mavrogeorgis E, Mischak H, Latosinska A, Siwy J, Jankowski V, Jankowski J. Reproducibility Evaluation of Urinary Peptide Detection Using CE-MS. Molecules. 2021;26(23).

20. Mischak H, Vlahou A, Ioannidis JP. Technical aspects and inter-laboratory variability in native peptide profiling: the CE-MS experience. Clin Biochem. 2013;46(6):432–43.

21. Mebazaa A, Casadio MC, Azoulay E, Guidet B, Jaber S, Levy B, et al. Post-ICU discharge and outcome: rationale and methods of the The French and euRopean Outcome reGistry in Intensive Care Units (FROG-ICU) observational study. BMC Anesthesiol. 2015;15:143.

22. Alkhalaf A, Zurbig P, Bakker SJ, Bilo HJ, Cerna M, Fischer C, et al. Multicentric validation of proteomic biomarkers in urine specific for diabetic nephropathy. PLoS One. 2010;5(10):e13421.

23. Delles C, Schiffer E, von Zur Muhlen C, Peter K, Rossing P, Parving HH, et al. Urinary proteomic diagnosis of coronary artery disease: identification and clinical validation in 623 individuals. J Hypertens. 2010;28(11):2316–22.

24. Frantzi M, van Kessel KE, Zwarthoff EC, Marquez M, Rava M, Malats N, et al. Development and Validation of Urine-based Peptide Biomarker Panels for Detecting Bladder Cancer in a Multi-center Study. Clin Cancer Res. 2016;22(16):4077–86.

25. He T, Melgarejo JD, Clark AL, Yu YL, Thijs L, Diez J, et al. Serum and urinary biomarkers of collagen type-I turnover predict prognosis in patients with heart failure. Clin Transl Med. 2021;11(1):e267.

26. He T, Mischak M, Clark AL, Campbell RT, Delles C, Diez J, et al. Urinary peptides in heart failure: a link to molecular pathophysiology. Eur J Heart Fail. 2021;23(11):1875–87.

27. Htun NM, Magliano DJ, Zhang ZY, Lyons J, Petit T, Nkuipou-Kenfack E, et al. Prediction of acute coronary syndromes by urinary proteome analysis. PLoS One. 2017;12(3):e0172036.

28. Huang QF, Trenson S, Zhang ZY, Yang WY, Van Aelst L, Nkuipou-Kenfack E, et al. Urinary Proteomics in Predicting Heart Transplantation Outcomes (uPROPHET)-Rationale and database description. PLoS One. 2017;12(9):e0184443.

29. Kuznetsova T, Mischak H, Mullen W, Staessen JA. Urinary proteome analysis in hypertensive patients with left ventricular diastolic dysfunction. Eur Heart J. 2012;33(18):2342–50.

30. Lindhardt M, Persson F, Zurbig P, Stalmach A, Mischak H, de Zeeuw D, et al. Urinary proteomics predict onset of microalbuminuria in normoalbuminuric type 2 diabetic patients, a sub-study of the DIRECT-Protect 2 study. Nephrol Dial Transplant. 2017;32(11):1866–73.

31. Packham DK, Wolfe R, Reutens AT, Berl T, Heerspink HL, Rohde R, et al. Sulodexide fails to demonstrate renoprotection in overt type 2 diabetic nephropathy. J Am Soc Nephrol. 2012;23(1):123–30.

32. Rossing K, Bosselmann HS, Gustafsson F, Zhang ZY, Gu YM, Kuznetsova T, et al. Urinary Proteomics Pilot Study for Biomarker Discovery and Diagnosis in Heart Failure with Reduced Ejection Fraction. PLoS One. 2016;11(6):e0157167.

33. Rotbain Curovic V, Magalhaes P, He T, Hansen TW, Mischak H, Rossing P, et al. Urinary peptidome and diabetic retinopathy in the DIRECT-Protect 1 and 2 trials. Diabet Med. 2021;38(9):e14634.

34. Rudnicki M, Siwy J, Wendt R, Lipphardt M, Koziolek MJ, Maixnerova D, et al. Urine proteomics for prediction of disease progression in patients with IgA nephropathy. Nephrol Dial Transplant. 2021;37(1):42–52.

35. Tofte N, Lindhardt M, Adamova K, Bakker SJL, Beige J, Beulens JWJ, et al. Early detection of diabetic kidney disease by urinary proteomics and subsequent intervention with spironolactone to delay progression (PRIORITY): a prospective observational study and embedded randomised placebo-controlled trial. Lancet Diabetes Endocrinol. 2020;8(4):301–12.

36. Verbeke F, Siwy J, Van Biesen W, Mischak H, Pletinck A, Schepers E, et al. The urinary proteomics classifier chronic kidney disease 273 predicts cardiovascular outcome in patients with chronic kidney disease. Nephrol Dial Transplant. 2021;36(5):811–8.

37. Zhang Z, Staessen JA, Thijs L, Gu Y, Liu Y, Jacobs L, et al. Left ventricular diastolic function in relation to the urinary proteome: a proof-of-concept study in a general population. Int J Cardiol. 2014;176(1):158–65.

38. Nkuipou-Kenfack E, Latosinska A, Yang WY, Fournier MC, Blet A, Mujaj B, et al. A novel urinary biomarker predicts 1-year mortality after discharge from intensive care. Crit Care. 2020;24(1):10.

39. Zhao X, Kwan JYY, Yip K, Liu PP, Liu FF. Targeting metabolic dysregulation for fibrosis therapy. Nat Rev Drug Discov. 2020;19(1):57–75.

40. Dweck MR, Joshi S, Murigu T, Alpendurada F, Jabbour A, Melina G, et al. Midwall fibrosis is an independent predictor of mortality in patients with aortic stenosis. J Am Coll Cardiol. 2011;58(12):1271–9.

41. Ekstedt M, Hagstrom H, Nasr P, Fredrikson M, Stal P, Kechagias S, et al. Fibrosis stage is the strongest predictor for disease-specific mortality in NAFLD after up to 33 years of follow-up. Hepatology. 2015;61(5):1547–54.

42. Gulati A, Jabbour A, Ismail TF, Guha K, Khwaja J, Raza S, et al. Association of fibrosis with mortality and sudden cardiac death in patients with nonischemic dilated cardiomyopathy. JAMA. 2013;309(9):896–908.

43. Currie GE, von Scholten BJ, Mary S, Flores Guerrero JL, Lindhardt M, Reinhard H, et al. Urinary proteomics for prediction of mortality in patients with type 2 diabetes and microalbuminuria. Cardiovasc Diabetol. 2018;17(1):50.

44. Batra R, Uni R, Akchurin OM, Alvarez-Mulett S, Gomez-Escobar LG, Patino E, et al. Urine-based multi-omic comparative analysis of COVID-19 and bacterial sepsis-induced ARDS. Mol Med. 2023;29(1):13.

45. Bannaga A, Metzger J, Voigtlander T, Pejchinovski M, Frantzi M, Book T, et al. Pathophysiological Implications of Urinary Peptides in Hepatocellular Carcinoma. Cancers (Basel). 2021;13(15).

46. Wei D, Melgarejo JD, Thijs L, Temmerman X, Vanassche T, Van Aelst L, et al. Urinary Proteomic Profile of Arterial Stiffness Is Associated With Mortality and Cardiovascular Outcomes. J Am Heart Assoc. 2022;11(8):e024769.

47. Piedrafita A, Siwy J, Klein J, Akkari A, Amaya-Garrido A, Mebazaa A, et al. A universal predictive and mechanistic urinary peptide signature in acute kidney injury. Crit Care. 2022;26(1):344.

